# Inhibitory control in children and adolescents with paediatric-onset obsessive-compulsive disorder - An fMRI study

**DOI:** 10.1101/2025.05.09.25327300

**Authors:** Kit Melissa Larsen, Valdemar Funch Uhre, Vytautas Labanauskas, Kristoffer Hougaard Madsen, Ayna Baladi Nejad, William Baaré, Kerstin Jessica Plessen, Hartwig Roman Siebner, Anne Katrine Pagsberg

**Author notes:** Corresponding author: Kit Melissa Larsen, Kettegård allé 36, 2650 Hvidovre. Equal contributions (Kit Melissa Larsen, Valdemar Funch Uhre). Equal contributions (Hartwig Roman Siebner, Anne Katrine Pagsberg).

## Abstract

**Background:** The pathophysiology of obsessive-compulsive disorder (OCD) appears to involve dysfunctions in brain circuits underlying sensorimotor, cognitive, affective, and motivational processes. One area of dysfunction observed is disruption in inhibitory motor control. While functional magnetic resonance imaging (fMRI) has revealed associated alterations at the brain network level in adults, studies in pediatric OCD has yielded inconsistent results.

**Methods:** This task-based fMRI study examined executive motor control in 65 unmedicated paediatric OCD patients and 58 age- and sex-matched healthy controls. Participants were aged eight to 17 years and performed a stop-signal task during whole-brain 3 Tesla fMRI. In addition to whole-brain analyses, we performed a region-of-interest analysis, focusing on brain regions subserving inhibitory motor control during the stop-signal task.

**Results:** The paediatric patients with OCD showed comparable task performance and inhibition abilities to healthy controls. During both successful and failed inhibition, patients with paediatric OCD showed comparable whole brain activation patterns as compared to healthy controls.

**Conclusion:** Our behavioural findings indicate typical inhibitory motor control proficiency in paediatric OCD patients together with comparable neural activation patterns when compared to controls. These findings, obtained in a large sample of unmedicated children and adolescents, suggest that alterations in inhibitory control may not be a prominent feature of early-stage OCD.

## 1. Introduction

Obsessive-compulsive disorder (OCD) is a debilitating disorder characterized by persistent and intrusive obsessions and repetitive compulsions (Association, 2013) that affects 2-3% of the population (Barton & Heyman, 2016; Canals, Hernández-Martínez, Cosi, & Voltas, 2012; Ruscio, Stein, Chiu, & Kessler, 2010). The onset of OCD commonly occurs in late adolescence (Anholt et al., 2014; Jagannathan, Ginger, Yu, Chasson, & Leventhal, 2024). However, previous research demonstrate differences between paediatric-onset and adult-onset OCD in clinical presentation, gender distribution, genetics, neurocognitive profile, and treatment response, indicating that paediatric-onset OCD constitutes a developmental subtype of OCD (Geller et al., 2001; Geller, Homayoun, & Johnson, 2021).

Obsessions and compulsions in OCD may be associated with deficits in inhibitory control, which refers to the reduced ability to suppress actions and thoughts when necessary (Chamberlain, Blackwell, Fineberg, Robbins, & Sahakian, 2005). Inhibitory motor control can be assessed in experiments either using interfering stimuli (e.g., Flanker task) or including a pre-potent response which occasionally must be either altered (e.g., switching task), withheld (e.g., Go/No-go task) or cancelled (e.g., stop signal task) (Aron, 2011; van Velzen, Vriend, de Wit, & van den Heuvel, 2014). A recent review of candidate neurocognitive endophenotypes in adult OCD suggests that impairment of inhibitory control may manifest only in adulthood (Marzuki, Pereira de Souza, Sahakian, & Robbins, 2020).

The inhibitory network associated with the stop-signal task includes left and right putamen and subthalamic nucleus (STN), together with left and right presupplementary motor area (pre-SMA) and the right inferior frontal gyrus (IFG) (Aron, 2011; Hannah & Aron, 2021; Swick, Ashley, & Turken, 2011; Verbruggen et al., 2019; Zhang, Geng, & Lee, 2017). The opercular part of the right IFG together with the right pre-SMA likely play a crucial role in the initial generation of the stop-command (Hannah & Aron, 2021) during inhibition. Recent meta-analytic evidence from functional neuroimaging studies of OCD indicates aberrant activation of cortico-striato-thalamo-cortical (CSTC) networks during the performance of inhibitory control tasks (Carlisi et al., 2017; Norman et al., 2016, 2019). This evidence is primarily based on findings from studies of adults with OCD and, thus, may not generalise to individuals with onset during childhood or adolescence. Paediatric-onset OCD often occurs during a vulnerable age period when the neural mechanisms supporting inhibitory control are not fully matured (Constantinidis & Luna, 2019; Madsen et al., 2020). Accordingly, age-related deviations in the development of inhibitory control networks warrant consideration when investigating OCD in paediatric and adolescent populations (Huyser, Veltman, de Haan, & Boer, 2009).

Functional magnetic resonance imaging (fMRI) studies on inhibitory control in children and adolescents with OCD have yielded conflicting results. One study found reduced interference-related activation during the multisource interference task (Fitzgerald et al., 2010) (n=18, age range 8-18 years), while others reported increased interference-related (Huyser, Veltman, Wolters, De Haan, & Boer, 2011) (n=25, age range 8-19 years) and set-shifting-related (Britton et al., 2010) (n=15, age range 10-17 years) activation in multiple regions within the CSTC network during a Flanker task and set-shifting task, respectively. Additional studies showed reduced interference-related, switching-related, and successful-inhibition-related activation of CSTC networks in a small group of boys (n=10, mean age 14.3 years) with remitted OCD, as compared to healthy controls, during the performance of several different inhibitory control tasks (Rubia et al., 2010; Rubia, Cubillo, Woolley, Brammer, & Smith, 2011; Woolley et al., 2008). Interestingly, a recent large study including 69 paediatric patients with OCD (8 through 19 years of age) and 72 healthy individuals found no group effects on interference-related activation in the multisource interference task (Fitzgerald et al., 2018). This finding is consistent with the absence of group differences in task-related activation during interference control in the multisource interference task (age range 8-18 years, n=21) (Fitzgerald et al., 2013) and during successful inhibition in the stop-signal task (age range 8-12 years, n=16) (Gooskens et al., 2019). As most studies examined medicated paediatric OCD patients, pharmacological effects might have confounded the observed results (Boedhoe et al., 2017, 2018). Consequently, it remains unclear whether, and to what extent, unmedicated children and adolescents with paediatric-onset OCD may display deviant brain activation patterns during inhibitory motor control tasks.

In this task-based fMRI study, we examined inhibitory control in paediatric OCD, addressing two main aims. First, we sought to identify alterations in brain activation during inhibitory motor control. Second, we explored how these changes relate to inhibitory performance and OCD symptom severity. Using the stop-signal task, we specifically focused on the capacity to withhold a prepotent motor response on infrequent stop-trials. We hypothesized that children with OCD will show impaired inhibitory control together with aberrant neural engagement of brain regions engaged in stopping.

## 2. Methods

### 2.1. Participants

A total of 130 children and adolescents with OCD and 90 healthy controls were enrolled in the TECTO trial between September 2018 and June 2022 (Pagsberg et al., 2022), which is a randomised clinical trial with a parallel case-control design comparing treatment effects of two family-based psychotherapies (ClinicalTrials.gov Identifier NCT03595098). A subset of 94 OCD patients and 82 healthy individuals were included in the present MRI sub-study (see Figure 1 for an overview). Patients were recruited from the Child and Adolescent Mental Health Center (CAMHC), Copenhagen University Hospital – Mental Health Services-CPH, Denmark. Inclusion criteria were 8 through 17 years of age, a primary diagnosis of OCD (F42 according to the criteria of the International Classification of Diseases, 10th revision, ICD-10) (WHO, 1992) and a Children’s Yale-Brown Obsessive Compulsive Scale (CY-BOCS) (Scahill et al., 1997) score of at least 16 (corresponding to moderate-to-severe OCD). Patients were excluded if they met ICD-10 criteria for a diagnosis of schizophrenia, psychosis, mania, bipolar disorder, substance dependence syndrome, or pervasive developmental disorder (not including Asperger’s syndrome). Additionally, patients were excluded if they had received treatment within the past six months using cognitive behavioural therapy, relaxation therapy, antidepressants, or antipsychotic medication. Healthy controls were recruited from the regional population via a random sample drawn from the Danish Central Person Registry (Pedersen, Heine, Møller, Ostrup, & Mortensen, 2011) that was matched on sex and age (within 3 months). Healthy controls were excluded if they met ICD-10 criteria for any current or previous psychiatric disorder. Both healthy participants and participants with OCD were excluded if they had a full-scale IQ below 70, had any neurological illness, or had previously sustained severe head trauma. Additional details on the TECTO trial are available in the trial protocol (Pagsberg et al., 2022). The study was approved by the National Research Ethics Committee (H-18010607) and The Knowledge Centre on Data Protection Compliance in The Capital Region of Denmark (VD-2018-263, l-suite 6502), and followed the Helsinki Declaration (“World Medical Association Declaration of Helsinki: ethical principles for medical research involving human subjects,” 2013). Participants provided signed informed consent (participants below the age of 15 years gave their assent and consent was provided by their guardian). Consent to partake in the MRI sub-study was not a prerequisite for participation in the TECTO trial.

**Figure 1.**
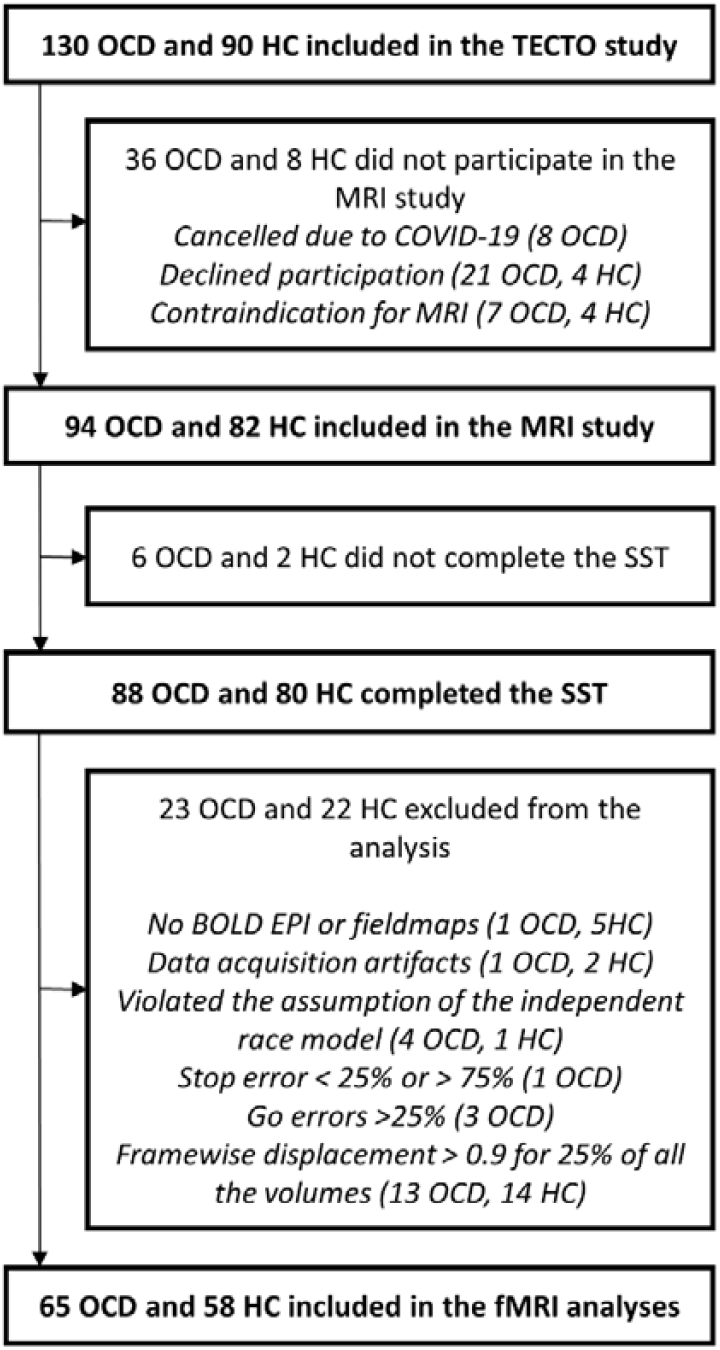
Participant flow diagram for the MRI study. Abbreviations: HC, healthy control; OCD, patients with obsessive-compulsive disorder; SST, stop signal task; MRI, magnetic resonance imaging; fMRI, functional magnetic resonance imaging; BOLD, Blood-Oxygen-Level Dependent; EPI, echo-planar MR imaging.

### 2.2. Clinical assessments

OCD diagnosis and psychiatric co-morbidities in patients and absence of any current or lifetime psychiatric disorders in healthy control was determined based on transference of item criteria from a semi-structured diagnostic interview-the Danish translation of the Kiddie Schedule for affective disorders and Schizophrenia – present and lifetime (K-SADS-PL) (Kaufman et al., 1997). OCD symptom severity was assessed with the CY-BOCS and illness severity was furthermore assessed by the Clinical Global Impression – Severity (CGI-S) (Busner & Targum, 2007). For both participant groups the level of psychosocial functioning was assessed with the Children’s Global Assessment Scale (C-GAS)(Shaffer et al., 1983). Participants’ handedness was evaluated via the 10-item version of the Edinburgh Handedness Inventory (EHI), employing a five-alternative forced-choice format (Oldfield, 1971) and summarized in a single laterality quotient. Additionally, motor coordination and speed were assessed with the Purdue Pegboard Test (PPBT)(Tiffin & Asher, 1948). The stage of puberty was assessed by self-report on the Tanner scale (Taylor et al., 2001). Finally, general cognitive ability (full-scale IQ) was assessed using age-appropriate versions of the Wechsler Intelligence Scales (WISC-V or WAIS-IV)(Wechsler, 2008).

### 2.3. Virtual-reality and Mock MRI

Some participants find MRI procedures discomforting, with discomfort often higher in children and adolescents and in clinical groups experiencing high levels of anxiety (Everts et al., 2021). In addition, head motion during brain MRI poses a major challenge in paediatric (Afacan et al., 2016) and psychiatric populations (Makowski, Lepage, & Evans, 2019). Participants experienced a narrated virtual reality (VR) simulation of the MRI procedure prior to the day of scanning. This simulation was specifically developed for the study to closely mimic the actual procedures. Additionally, participants underwent a mock scan procedure immediately before the actual scan (Model PST-100355, Psychology Software Tools, Inc.). The mock scanner is a full-scale MRI machine replica without internal magnets and was used to practice the fMRI task, lying still while receiving visual live feedback on head movements and familiarize the participant with the scan environment and gradient sounds.

### 2.4. Stop signal task

The stop signal task was presented with PsychoPy [version 1.84.2] and consisted of a total of 220 trials, including 160 go-trials, 40 stop-trials, and 20 null-trials (Figure 2). Go-trials and stop-trials were presented in pseudo-randomized orders, with the constraints that the first 12 trials were go-trials and that a stop-trial could not be followed by another stop-trial. The go-trials were presented with either an upward or downward pointing arrow (Figure 2), indicating if participants were to respond with their right or left index finger, respectively. The mapping between arrow direction and response hand (i.e., whether the upward arrow indicated the right or left finger) was counterbalanced across participants. Participants were instructed to respond as quickly and accurately as possible to the direction of the centrally presented arrow unless interrupted by a red circle - the stop signal - after a variable stop-signal delay (SSD; stop-trial), in which participants had to refrain from responding. We used a linear staircase algorithm to adaptively adjust task difficulty in increments of 50 ms by increasing the SSD after successful stop-trials to make the task harder or decreasing it after failed stop-trials to make the task easier. The initial SSD was subject-specific and was determined by averaging the SSDs of the last half of the mock session’s stop trials and rounding to the nearest 50 ms. The initial SSD used in the mock session was 250 ms. Using a subject-specific initial SSD during the fMRI task ensures that the SSD distribution is adequately sampled around a participant’s mean SSD during task performance. Participants were post hoc excluded from further analysis if more than 25% of go-trials resulted in errors, if the error rate on stop trials was outside the 25-75% range (Congdon et al., 2012), or if the assumption of independence of go and stop processes was violated (i.e., if the mean reaction time (RT) on failed stop-trials was greater than the mean RT on go-trials). For the remaining participants, we calculated the stop-signal reaction time (SSRT) using the non-parametric integration method with replacement of go omission errors (Verbruggen et al., 2019). The SSRT is the estimated time it takes to cancel a prepotent motor action successfully. A shorter SSRT reflects faster motor response inhibition, while a longer SSRT indicates slower cancellation of initiated motor responses.

**Figure 2.**
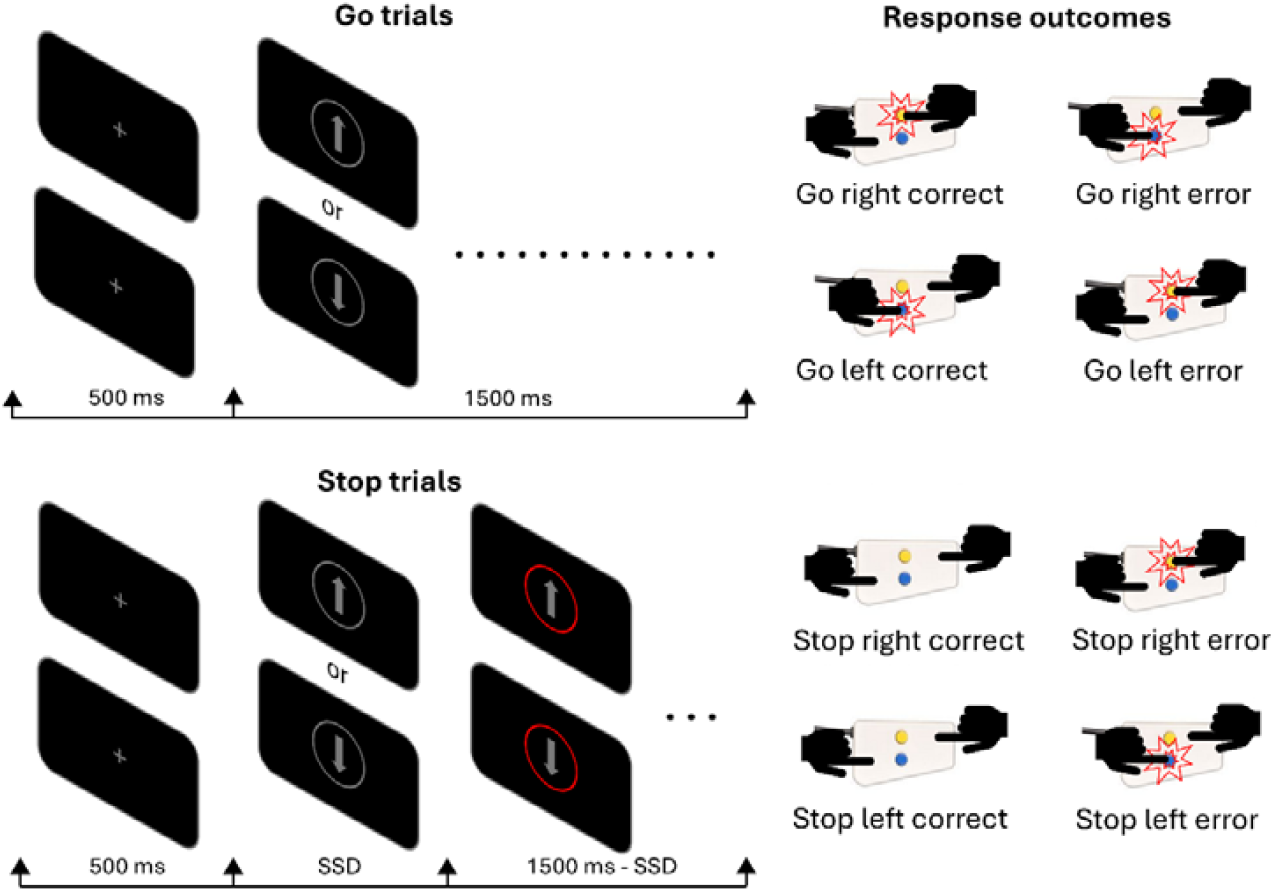
Trial outline of the stop signal task. Participants completed 220 trials (160 go-trials, 40 stop-trials and 20 null-trials). The first 12 trials were go-trials, and a stop-trial could not be followed by another stop-trial. Each trial started with a fixation cross presented for 500 ms. Participants were instructed to respond as quickly and accurately as possible to the direction of a centrally presented arrow pointing upward or downward (go-trials) unless it was interrupted by a red circle (stop signal) after a variable stop-signal delay (SSD, stop-trials) where participants had to refrain from responding. The response device was placed on the chest of the participant. The response mapping was counterbalanced across participants: half of the participants were instructed to press the upper button with their right hand and the lower button with their left hand, while the other half had the reverse configuration. Go trials were classified as correct go (left or right) or go error: a commission error (incorrect button press) or omission error (no button press). Stop trials were classified as stop error (left or right) or stop correct (left, or right, no button press). Additionally, we included null trials, which occurred on every 11^th^ trial (except the first round of 12 go-trials) and consisted of a prolonged presentation of the fixation cross, matching the duration of a normal trial.

### 2.5. Image acquisition

Whole-brain MRI was performed with a 3T Philips Achieva scanner and a 32-channel head coil at the Danish Research Centre for Magnetic Resonance (DRCMR). For each participant, a total of 530 single-shot gradient-echo echo-planar images (EPI; TR=1.700 s, TE=0.03s, FA=71, matrix size=68×70 mm, 32 slices in the axial orientation, slice excitation in ascending order, slice thickness=3.4 mm, slice gap=0.6 mm, FOV=120mm) were acquired in addition to another five EPIs with opposing phase encoding direction. In addition, a T1-weighted image was acquired for each participant (matrix size= 288×288 mm, voxel size= 1 mm isotropic, TE= 2.71ms, TR=5.98ms, FA=8, 245 slices in the sagittal orientation) with navigator-based prospective motion correction to improve image quality (Andersen, Björkman-Burtscher, Marsman, Petersen, & Boer, 2019). Foam pads were used to reduce head motion during scanning. Participants were shown a calming movie of fish and turtles during the structural scan to reduce head motion (Greene et al., 2018). Weighted blankets were available to help reduce movement and increase comfort.

### 2.6. Pre-processing of the fMRI data

MRI data were pre-processed using default parameters of fMRIPrep (version 24.0.1) (Esteban et al., 2019). T1w images were corrected for intensity non-uniformity and skull-stripped. Brain tissue segmentation of cerebrospinal fluid, white-matter and gray-matter was performed on the brain-extracted T1w and used to create subject-specific brain masks. Volume-based spatial normalization through non-linear registration (ANTs 2.5.1) was performed using the 4.5 to 18.5 years age range MNIPediatricAsym:cohort-1 template from TemplateFlow (Ciric et al., 2022). Functional EPI images were slice time corrected and realigned to the time series mean. Fieldmap images obtained with opposite phase encoding direction were used to estimate susceptibility distortions (PEB (phase-encoding based) and PEPOLAR (phase-encoding polarity)). The corrected BOLD images were then co-registered to the T1w reference image. Finally, the spatially normalised BOLD images were smoothed with an isotropic 6 mm full width at half maximum (FWHM) Gaussian kernel.

### 2.7. Statistical analyses

#### 2.7.1. Bayesian inference

To assess if the groups were different with respect to the behavioural and demographic variables, we used Bayesian analyses performed with R (4.2.2) and in Rstudio (version 2020.07.2) using the package BayesFactor (version 0.9.12-4.7) (Morey, R.D. and Rouder, 2022). The appropriate Bayesian Student’s t-tests and Bayesian Contingency Tables were used for the demographics data. To assess group differences in the behavioural data, Bayesian linear regression was used with age and sex as covariates. Levels of evidence are reported according to the standard interpretation of Bayes factors (BF) in favour of the alternative hypothesis, i.e., patients with OCD differ from healthy controls (BF_10_). A BF of 1 represents no evidence, 1-3 anecdotal evidence, 3-10 moderate evidence, 10-30 strong evidence, 30-100 very strong evidence, and >100 decisive evidence (van Doorn et al., 2020).

#### 2.7.2. First-level GLM

At the first level GLM, we modelled correct left- and right-hand go-trials, correct left and right stop trials and failed left and right stop trials using a stick function at the onsets of the trial cues. We additionally included combined left- and right-hand go errors as a separate regressor. Null trials were not modelled and constituted an implicit baseline. The regressors of no interest included framewise displacement (FD; weighted average of rotational and translational displacements), six rigid-body motion parameters (translation and rotation) and principal components capturing physiological noise derived using the data-driven anatomical Component-Based Noise Correction (aCompCor) method (Behzadi, Restom, Liau, & Liu, 2007). Furthermore, the analysis incorporated a high pass filter with a 128 Hz cut-off frequency. The analysis was carried out using nilearn (version 0.10.4) (Abraham et al., 2014). Participants having more than 25% of volumes censored were excluded from all further analyses to ensure sufficient data for statistical inference. Despite our efforts to mitigate scan-related discomfort and head movements by using VR and a Mock scan session, we had significant data loss due to participant movement during fMRI, resulting in the exclusion of 13 OCD patients and 14 healthy controls as depicted in Figure 1.

#### 2.7.3. Group-level whole-brain analyses

Contrast images for correct stop-trials versus correct go-trials (successful inhibition) and failed stop-trials versus correct go-trials (failed inhibition) were entered into separate second-level models to test for group differences in the activation patterns for failed and successful inhibition. Both models included mean FD, age, and sex as covariates. To ensure that the task evoked the expected activation pattern, we further looked at the activity associated with go-correct, stop-correct and failed-stop against this implicit baseline for both patients and controls, left and right-hand responses separately. For all analyses, we used non-parametric permutation inference, which does not rely on the usual parametric statistical assumptions, by employing the permutation analysis of linear models (PALM) tool (Winkler, Ridgway, Webster, Smith, & Nichols, 2014), with Threshold-Free Cluster Enhancement (TFCE) (Spisák et al., 2019). The analysis included 5000 permutations, and we tested for bidirectional effects. Results were considered significant at a familywise error corrected (FWE) threshold of p<.05.

#### 2.7.4. Region-of-interest analyses

We performed planned post-hoc region-of-interest (ROI) analyses within the well-documented network implicated in stopping when performing the stop-signal task (Hannah & Aron, 2021; Swick et al., 2011; Zhang et al., 2017), including left and right putamen, subthalamic nucleus (STN), and the right inferior frontal gyrus (IFG), defined using the PickAtlas toolbox (Maldjian, Laurienti, & Burdette, 2004; Maldjian, Laurienti, Kraft, & Burdette, 2003), left and right presupplementary motor areas (pre-SMA) as defined in (Neubert, Mars, Sallet, & Rushworth, 2015). Mean contrast estimates within the regions were extracted using nilearn from each participant during both failed inhibition and successful inhibition.

Group comparison of ROI activation was performed with Bayesian repeated-measures ANCOVAs with group (OCD patients vs. healthy controls) as a between-subject factor and ROIs (left and right putamen, STN and pre-SMA, and right IFG) as a within-subject factor. As with all other models, mean FD, age and sex were added as covariates.

As mentioned in the introduction of the paper, the opercular part of the right IFG together with the right pre-SMA likely play a crucial role in the initial generation of the stop-command (Hannah & Aron, 2021) during inhibition. Therefore, we explored whether activity within these regions of interest was related to SSRT during successful inhibition and whether the activation of these regions was associated with OCD symptom severity during either successful inhibition or failed inhibition. These analyses were performed using Bayesian partial correlation.

## 3. Results

### 3.1. Sample characteristics

Patients and controls did not differ in the proportion of females (BF_10_=0.570), stage of puberty (Tanner BF_10_=0.230), and motor coordination and speed as assessed with the PPBT left hand, right hand, and both hands (BF_10_ between 0.194 and 1.488), see Table 1 for an overview. Groups were also comparable in age (BF_10_=0.445), handedness (EHI laterality quotient BF_10_=0.288), and general cognitive ability (full-scale IQ BF_10_=1.030). There was decisive evidence for a group difference in parental education level (BF_10_=1032.490). Parents of healthy controls had a higher educational level.

**Table 1.** Demographics and clinical characteristics of the participants included in the fMRI analyses.

| Variable | OCD patients<br>(n=65)<br>Mean (SD) or n (%) | Healthy controls<br>(n=58)<br>Mean (SD) or n (%) | $BF_{10}$ |
| --- | --- | --- | --- |
| **Demographic measures** |  |  |  |
| Female | 33 (50.7%) | 35 (60.3%) | 0.570 |
| Age in years | 14.14 (2.6)<br>Range: 8-17 | 13.5 (2.55)<br>Range: 8-17 | 0.445 |
| Tanner stage | 3.12 (1.33)<br>Range: 1-5 | 2.96 (1.29)<br>Range: 1-5 | 0.230 |
| EHI laterality quotient | 60.87 (74.78) | 72.72 (62.99) | 0.288 |
| Purdue Pegboard scores |  |  |  |
| Right hand | 14.37 (2.05) | 14.72 (2.21) | 0.278 |
| Left hand | 13.69 (1.61) | 13.67 (2.30) | 0.194 |
| Both hands | 11.69 (1.47) | 11.44 (2.16) | 0.247 |
| Assembly | 35.17 (7.69) | 38.31 (8.35) | 1.488 |
| Full-scale IQ (WISC-V/WAIS-IV) | 100.23 (13.14) | 105.08 (14.37) | 1.030 |
| Parental education level in years | 15.31 (2.08) | 17.01 (1.70) | 1032.49 |
| **Clinical measures** |  |  |  |
| CY-BOCS total score | 24.89(4.78) |  |  |
| *Obsessions* | 12.44 (2.51) |  |  |
| *Compulsions* | 12.44 (2.68) |  |  |
| C-GAS | 56.27 (9.26) | 89.3 (8.13) | 1.12e+36 |
| CGI-S | 4.55 (0.85) |  |  |
| Total with at least one co-occurring disorder | 35 (53.8%) |  |  |
| *Phobic anxiety disorders (F40)* | 1 (1.5%) |  |  |
| *Other anxiety disorders (F41)* | 2 (3%) |  |  |
| *Stress-related disorders (F43)* | 7 (10.7%) |  |  |
| *Eating disorders (F50)* | 3 (4.6%) |  |  |
| *Personality disorders (F61)* | 1 (1.5%) |  |  |
| *Pervasive developmental disorders (F84)\** | 9 (13.8%) |  |  |
| *Other developmental disorders (F88)* | 3 (4.6%) |  |  |
| *Attention-deficit hyperactivity disorders (F90)* | 6 (9.2%) |  |  |
| *Conduct disorders (F91)* | 1 (1.5%) |  |  |
| *Emotional disorders with childhood onset (F93)* | 4 (6.1%) |  |  |
| *Tic disorders (F95)* | 7 (10.7%) |  |  |
| *Other behavioral and emotional disorders with childhood onset (F98)* | 1 (1.5%) |  |  |

Note: OCD patients and healthy controls were compared pairwise using the Bayesian students t-test and Bayesian contingency tables. Data on the EHI was missing for four patients and two healthy controls. Abbreviations: BF<sub>10</sub>, Bayes Factor in favour of the alternative hypothesis (i.e., OCD patients differ from healthy controls) over the null hypothesis (OCD patients do not differ from healthy controls). BF=1: no evidence, BF=1-3: anecdotal evidence, BF=3-10: moderate evidence, BF=10-30: strong evidence, BF=30-100: very strong evidence, BF>100: decisive evidence; C-GAS, Children's Global Assessment Scale; CGI-S, Clinical Global Impression - Severity; CY-BOCS, Children's Yale-Brown Obsessive Compulsive Scale; EHI, Edinburgh Handedness Inventory; IQ, Intelligence Quotient; OCD, Obsessive-compulsive disorder; SD, standard deviation; TOCS, Toronto Obsessive-Compulsive Scale; WAIS-IV, Wechsler Adult Intelligence Scale version IV; WISC-V, Wechsler Intelligence Scale for Children version V. \* All cases were Asperger's syndrome (F84).

The OCD group showed a reduced global level of functioning compared to the healthy control group (C-GAS BF_10_=1.12e+36). In the OCD group, 30 of 65 (46.2%) had an exclusive diagnosis of OCD, while 35 of 65 (53.8%) had either one or two comorbid psychiatric disorders. The most common comorbid disorders were Asperger’s syndrome (13.8%), attention-deficit hyperactivity disorders (ADHD; 9.2%), and tic disorders (10.7%). On average, OCD patients had moderate to severe OCD symptoms (mean CY-BOCS total score of 24.89, SD=4.78), and illness severity was moderate to severe (mean CGI-S score of 4.55 SD=0.85).

### 3.2. No group differences in the behavioural outcomes of the stop signal task

During fMRI, patients and controls showed comparable task performance (Fig.3). There was anecdotal evidence against group differences regarding the RT for correct go-trial RT (ms), coefficient of variation (CV, %) in go-trial RT, SSRT (ms), go-trial omission errors (%), go-trial commission errors (%), and mean SSD (ms) (Bayes factors all between 0.191 and 0.293), see Figure 3 for distribution of data in the two groups and Table 2 for statistical comparisons.

**Table 2.** Stop signal task performance of the participants included in fMRI analyses. Abbreviations: BF_10_, Bayes Factor in favour of the alternative hypothesis (OCD patients differ from healthy participants) versus the null hypothesis (OCD patients do not differ from healthy controls); CV, coefficient of variation (SD/mean); OCD, Obsessive-compulsive disorder; RT, reaction time; SD, standard deviation; SSD, stop-signal delay; SSRT, stop-signal reaction time.

| | OCD n=65<br>Mean (SD) | Healthy controls n=58<br>Mean (SD) | $BF_{10}$ |
| --- | --- | --- | --- |
| CV in go-trial RT (%) | 24.16 (4.63) | 24.12 (4.27) | 0.201 |
| Mean correct go-trial RT (ms) | 615.96 (117.34) | 624.39 (110.61) | 0.191 |
| SSRT (ms) | 239.82 (54.71) | 230.89 (60.63) | 0.293 |
| Go-trial omission error (%) | 0.93 (1.70) | 1.19 (1.94) | 0.217 |
| Go-trial commission error (%) | 1.33 (1.76) | 1.19 (1.33) | 0.209 |
| SSD (ms) | 343.73 (124.02) | 360.67 (119.33) | 0.198 |

**Figure 3.**
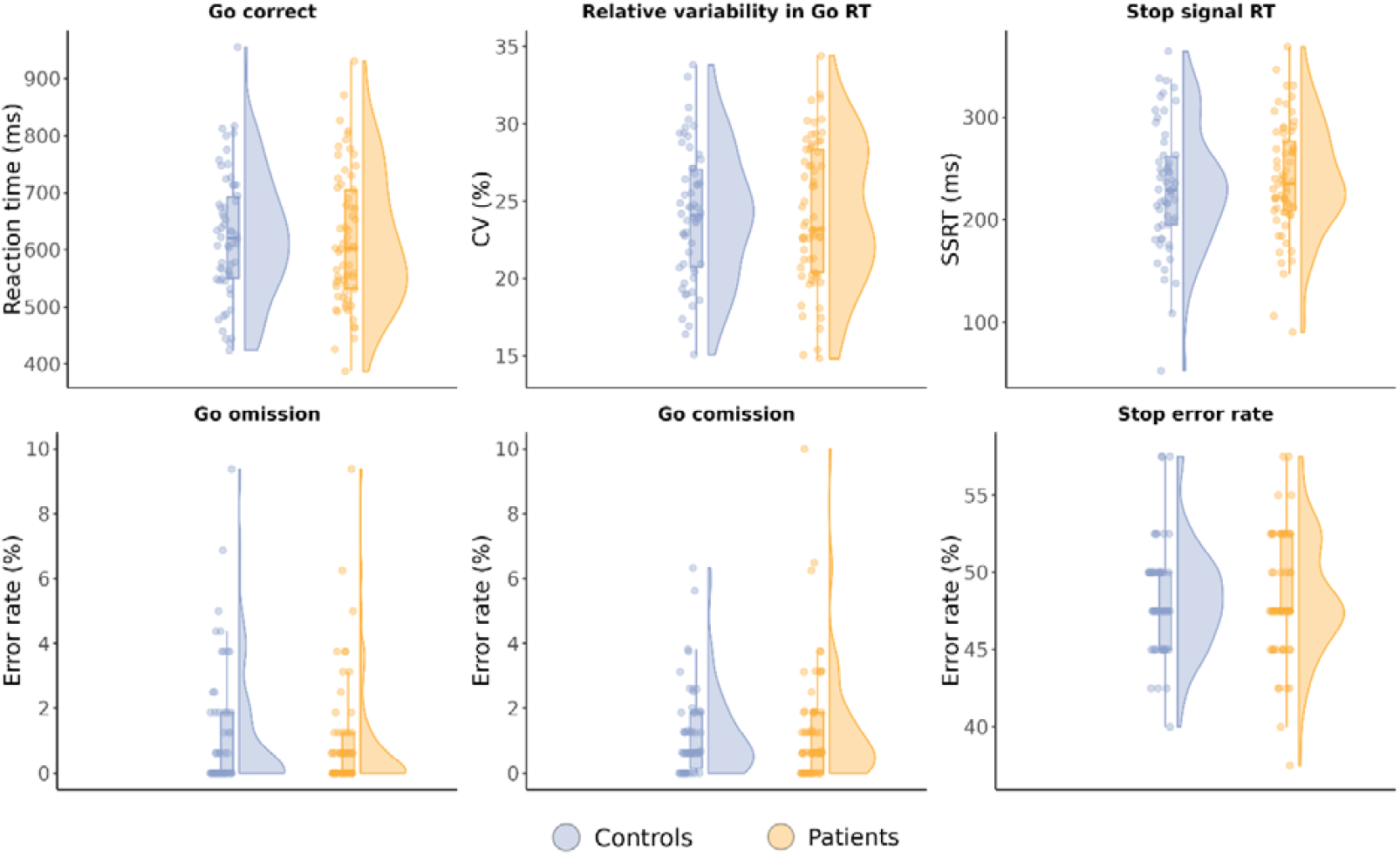
Performance of the stop signal task during fMRI for the OCD patients (light orange) and healthy controls (light blue) groups. The individual data points are shown on top of the box plots. We found moderate evidence that OCD patients and healthy controls showed comparable performance on mean go-trial RT (top left), CV in mean go-trial reaction time variability (top middle), stop signal reaction time (SSRT, top left), percentage of go-trial omission errors (bottom left), percentage of go-trial commission errors (bottom middle), and stop error rate (bottom right). Abbreviations: CV, coefficient of variation (SD/mean); RT, reaction time; OCD, obsessive-compulsive disorder. We used default settings to plot the density estimates, these are shown for visualization purposes.

### 3.3 Task effect of the stop signal task

To ensure that our task engaged the relevant brain areas associated with go and stopping, we investigated the main activity related to both go-correct, as well as successful and unsuccessful stop, see Figure S1. For the go-correct trials, both groups showed activation in the right and left primary motor cortex, postcentral gyri, right and left visual cortex as well as left insula, and cerebellum lobule VI. Controls further showed activity in the right putamen, and right and left thalamus. For the stop trials similar areas showed up, with additional activation in the hippocampus, parietal brain regions and supplementary motor area, extending to the anterior cingulate cortex during failed stop trials. Controls also showed activation in the subthalamic nucleus for both failed and successful stop as well as in dorsal anterior cingulate cortex for failed stop trials.

### 3.4. Children and adolescents with paediatric onset OCD show similar activation patterns during successful inhibition and failed inhibition

We then tested whether the group with paediatric OCD and the control group differed in brain activation patterns during both successful and failed inhibition. For this, we isolated the inhibitory activity by contrasting against the correct go responses (successful inhibition: stop correct < go correct and failed inhibition: stop error < go correct). During successful inhibition and failed inhibition there were no observed group differences.

In the post hoc ROI-analysis within the pre-defined inhibitory network comprising left and right putamen, STN, pre-SMA and the right IFG (Figure S2 and table S1), there were no differences in brain activity between patients and controls during either successful (stop correct > go correct) or failed inhibition (stop error > go correct). All Bayes factors were below 3.

We further explored whether activity within the right IFG and right pre-SMA were related to SSRT during successful inhibition, and whether the activation of these regions was associated with OCD symptom severity during either successful inhibition or failed inhibition (see Figure S3 and S4). We found no evidence for any of the explored associations (all Bayes factors were below 3, as indicated in the figures).

## 4. Discussion

We used task-based whole-brain fMRI to examine the functional brain network underlying the ability to inhibit a prepotent motor response in a large sample of unmedicated paediatric OCD patients and age- and sex-matched healthy controls. During both successful inhibition and failed inhibition, the paediatric OCD group showed comparable activation. Consistent with our findings, Gooskens et. al, 2021 reported no whole brain between-group differences in brain activity in a pediatric sample of OCD compared to healthy controls. However, Fitzgerald et. al, 2010 found increased activation in dorsal anterior cingulate in a smaller sample of 18 children with OCD when compared to controls during a multisource interference task (Fitzgerald et al., 2010). Further, a recent meta-analysis on inhibitory control found abnormal activation of the dorsal anterior cingulate cortex in patients with OCD compared to healthy controls. However, only three articles out of 21 included paediatric samples in that meta-analyses (Funch Uhre et al., 2022). Our results, therefore, extend the previous literature showing that the neural activation during both failed and successful inhibition is comparable to controls, in a large sample of unmedicated paediatric patients with OCD.

A limitation of studies within inhibitory control in paediatric OCD and OCD in general, is the inclusion of medicated participants. For example, a recent large fMRI study in children with OCD (Fitzgerald et al., 2018) yielded no significant group differences in brain activation during a multisource interference task. However, 34 out of 69 of the OCD patients underwent pharmacological treatment, predominantly with selective serotonin reuptake inhibitors (SSRIs). SSRIs have been shown to have a beneficial effect on the integrity of CSTC networks involved in inhibitory control (Niendam et al., 2012; Saxena et al., 2002). One previous study in an unmedicated cohort of 25 paediatric OCD patients, though, showed interference-related hypoactivation of the left dorsolateral prefrontal cortex and right insula during a Flanker task (Huyser et al., 2011). Overall, our findings significantly extend previous work and underscore the importance of considering medication status in neuroimaging studies of paediatric OCD.

Our finding that the patients with paediatric OCD and healthy controls demonstrated comparable task performance in the stop signal task fits previous behavioural results from fMRI studies using this task in paediatric OCD (Gooskens et al., 2019; Rubia et al., 2010; Woolley et al., 2008). Notably, these previous studies involved patients treated with SSRIs, which have shown to influence neural activity in the motor cortex (Gerdelat-Mas et al., 2005) and reduce the SSRT (Skandali et al., 2018) in adults. Since our study involved only unmedicated patients, comparable SSRT cannot be attributed to pharmacotherapy, but may rather suggest an intact motor inhibition in paediatric onset OCD. In addition, our finding is consistent with a recent meta-analysis indicating only a minimal deficit in SSRT across all ages in OCD, particularly in younger participants (Mar, Townes, Pechlivanoglou, Arnold, & Schachar, 2022), although again, the meta-analysis included a mix of medicated and non-medicated patients. Future longitudinal case-control studies using the stop signal task are needed to discern whether a prolongation of SSRT emerge with the progression of the disorder or represent a specific feature of adult-onset OCD. Likewise, prospective investigations are needed to determine if SSRT improves with reduced symptom severity in patients undergoing treatment.

Our study has several strengths. Our relatively large sample size gave us increased statistical power compared to previous smaller studies to detect true effects and reduced the likelihood that any observed effects were spurious findings (Button et al., 2013). We included moderate to severely ill paediatric OCD patients with significant functional impairments when compared to healthy controls, who were carefully assessed with the same validated diagnostic interview as the patients to ensure the absence of any prior and current psychiatric disorders. We carefully ensured the matching of age between groups and included age as a covariate in all our analyses. Another strength is the lack of confounding medication effects, since all patients were unmedicated.

Despite our efforts to mitigate scan-related discomfort and head movements by using VR and a Mock scan session, we had significant data loss due to participant movement during fMRI. However, this affected both groups equally. Also, even though the presence of comorbid disorders enhances the generalisability of findings since psychiatric comorbidities are very common in paediatric OCD (Sharma et al., 2021), it might have affected task performance as well as corresponding neural correlates in the OCD group (Arnold, Ickowicz, Chen, & Schachar, 2005), albeit the same cohort showed deficits in working memory, processing speed and cognitive flexibility (Uhre et al., 2023). Finally, we used an adaptive algorithm to individually adjust the SSD to secure a comparable level of task performance across participants. This introduces differences in task difficulty among participants (i.e., differences in SSD) that may have influenced the estimation of individual differences in inhibition-related brain activity (D’Alberto et al., 2018). Nevertheless, since both groups showed similar average SSDs, it seems unlikely that observed group differences in inhibition-related activity were influenced by group differences in objective task difficulty.

The present study presents baseline findings from the longitudinal interventional randomised clinical TECTO trial (Pagsberg et al., 2022). The next step will be to compare the effects of cognitive-behavioural therapy versus psychoeducation and relaxation therapy on neural and behavioural correlates of inhibitory control at follow up. To conclude, our findings suggest that neural correlates of inhibitory control in paediatric-onset OCD are not affected at the group level, neither the behaviour itself. This finding, obtained in a relatively large unmedicated sample, makes an important contribution to the understanding of paediatric-onset OCD and the development of response inhibition.

## Data Availability

Data is not publicly available, but can be retrieved during collaborations.

## Acknowledgement and funding

This study was funded by the Lundbeck Foundation, Capital Region Mental Health Services Research Foundation, Gangsted Foundation, Psychiatric Research Foundation of 1967, Holm’s Memorial Scholarship, Doctor Sofus Carl Emil Friis and wife Olga Friis’ Scholarship, Network for Research and Quality Assurance in Psychotherapy, Rosalie Petersen’s Foundation, Independent Research Fund Denmark.

HRS is supported by a Grand Solutions grant from Innovation Fund Denmark (9068-00025B) and a grant from the Lundbeck Foundation (grant nr. R336-2020-1035). SF was supported by the William Demant foundation. SF and HRS were supported by a grant from Novo Nordisk foundation (NNF17OC0027872). KML received funding from the Lundbeck Foundation (R322-2019-2311).

We thank all former and current members of the TECTO team for their substantial help with the recruitment and clinical assessment of study participants: Sofie Heidenheim Christensen, Julie Hagstrøm, Nicole Nadine Lønfeldt, Camilla Funch Uhre, Linea Pretzmann, Christine Thoustrup, Iben Clemmesen, Tin Aaen Gudmandsen, Nicoline Korsbjerg, Anna-Rosa Cecilie Mora-Jensen, Melanie Ritter, Emilie Thorsen, Klara Sofie Vangstrup Halberg, Merete Lindahl and Katrine Holmegaard Sørensen. We thank the Copenhagen Trial Unit for managing the electronic case report forms. We thank Sussi Larsen, Frederik Espensen, Vincent Boer, Kevin Pedersen, Christian Bauer and Daban Sulaiman for their help with the acquisition of imaging data. Valdemar Funch Uhre and Ayna Baladi Nejad changed employment to Novo Nordisk A/S.

## CRediT author statement

**Kit Melissa Larsen**: Methodology, Software, Data analysis and interpretation of results, Writing – Review & Editing, Supervision. **Valdemar Funch Uhre**: Conceptualization, Methodology, Data analysis and Interpretation of Results, Software, Formal analysis, Investigation, Data curation, Writing – First draft, Writing – Review & Editing, Visualization. **Vytautas Labanauskas**: Data analysis and interpretation of results, Review and Editing. **Kristoffer Hougaard Madsen**: Software, Resources, Writing – Review & Editing. **Ayna Baladi Nejad**: Methodology, Software, Resources, Writing – Review & Editing, Supervision. **William Baaré**: Methodology, Software, Resources, Writing – Review & Editing, Supervision. Kerstin Jessica Plessen: Conceptualization, Methodology, Writing – Review & Editing, Supervision, Project administration, Funding acquisition. **Hartwig Roman Siebner**: Conceptualization, Methodology, Data analysis and interpretation of results, Writing – Review & Editing, Supervision, Project administration, Funding acquisition. **Anne Katrine Pagsberg**: Conceptualization, Methodology, Data analysis and interpretation of results, Writing – Review & Editing, Supervision, Project administration, Funding acquisition.

