## Supplementary material for "Inhibitory control in children and adolescents with paediatric-onset obsessive-compulsive disorder - An fMRI study"

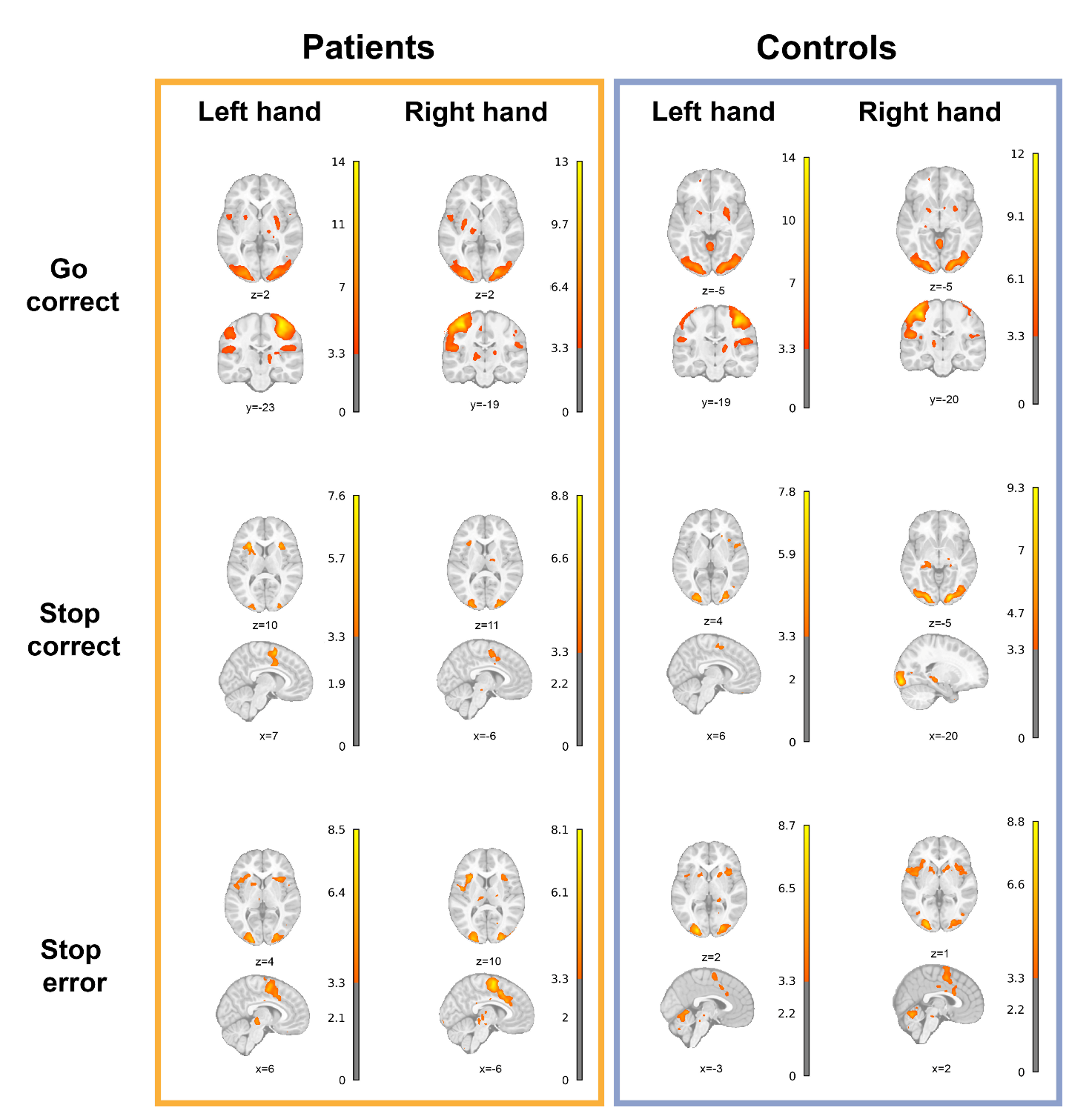

*Figure S1. Stop signal task related activations and deactivations in patients (n=65) and controls (n=58), during correct go (Go correct>0, left- and right-hand responses), correct stop (Stop correct>0, left- and right-hand stops) and incorrect stop trials (Stop error>0, left or right cue error). The BOLD images are uncorrected p<.001. The colour bar represents t-stat values. A t-value of 3.3 equals p<0.001 uncorrected. Brain images are shown using neurological convention (left side of the brain is on the left side of the image, right side of the brain on the right side of the image).*

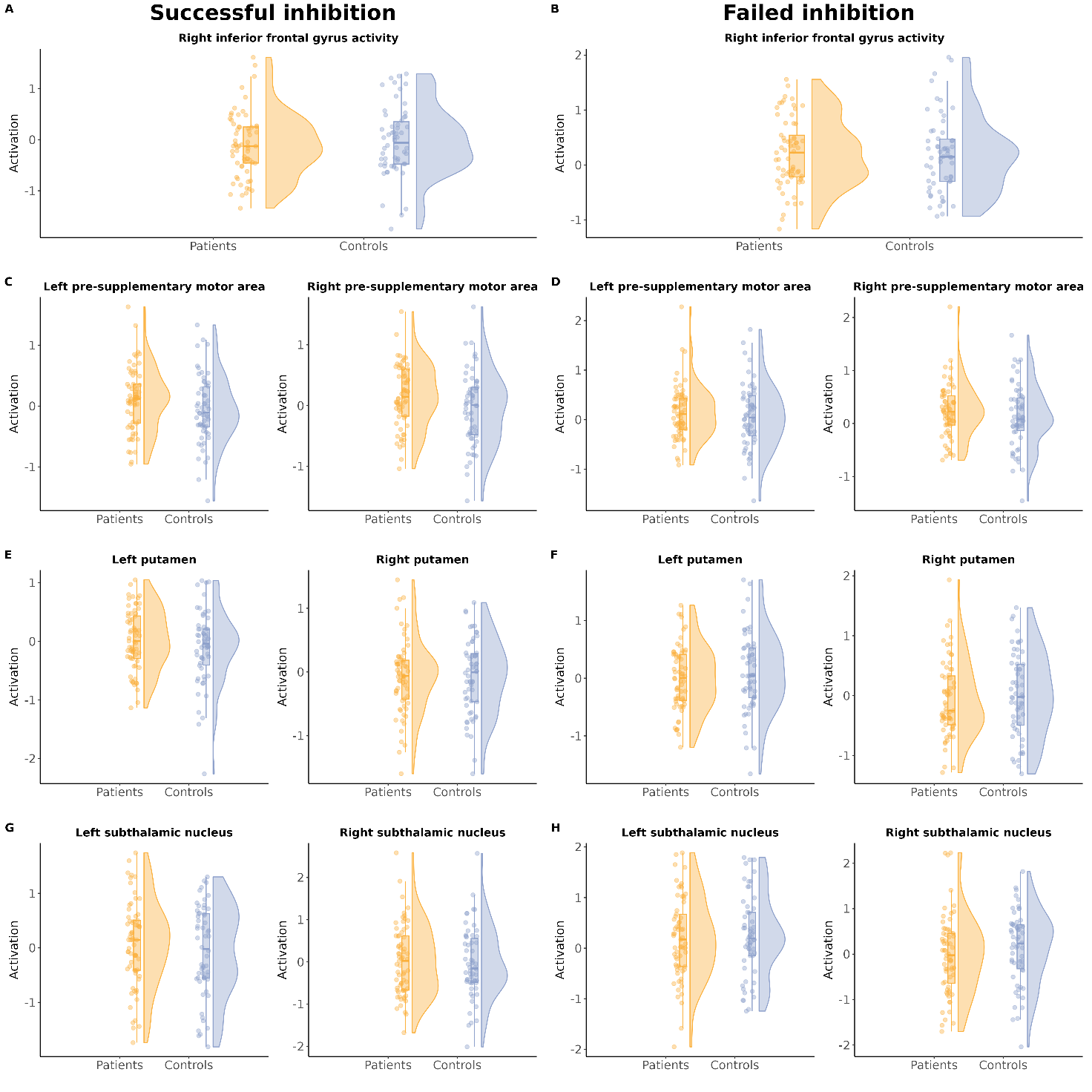

Figure S2: Activity in the regions of interest during successful inhibition. Blue colours represent controls, and orange colours represent patients. The regions of interest are: Left and right subthalamic nucleus (STN), left and right putamen, left and right pre-supplementary motor area and right inferior fontal gyrus.

*Table S1: Bayes factors indicating the evidence for a group difference in the seven regions of interest, during both successful inhibition and failed inhibition. BF_10_, Bayes Factor in favour of the alternative hypothesis (OCD patients differ from healthy controls) versus the null hypothesis (OCD patients do not differ from healthy controls).*

|  | **Successful inhibition** | **Failed inhibition** |
| --- | --- | --- |
| **Regions of interest** | **BF_10_** | **BF_10_** |
| Left putamen | 0.063 | 0.008 |
| Right putamen | 1.957 | 0.006 |
| Left STN | 0.021 | 0.021 |
| Right STN | 0.004 | 0.008 |
| Left pre-SMA | 0.037 | 0.007 |
| Right pre-SMA | 0.109 | 0.014 |
| Right IFG | 0.111 | 0.731 |

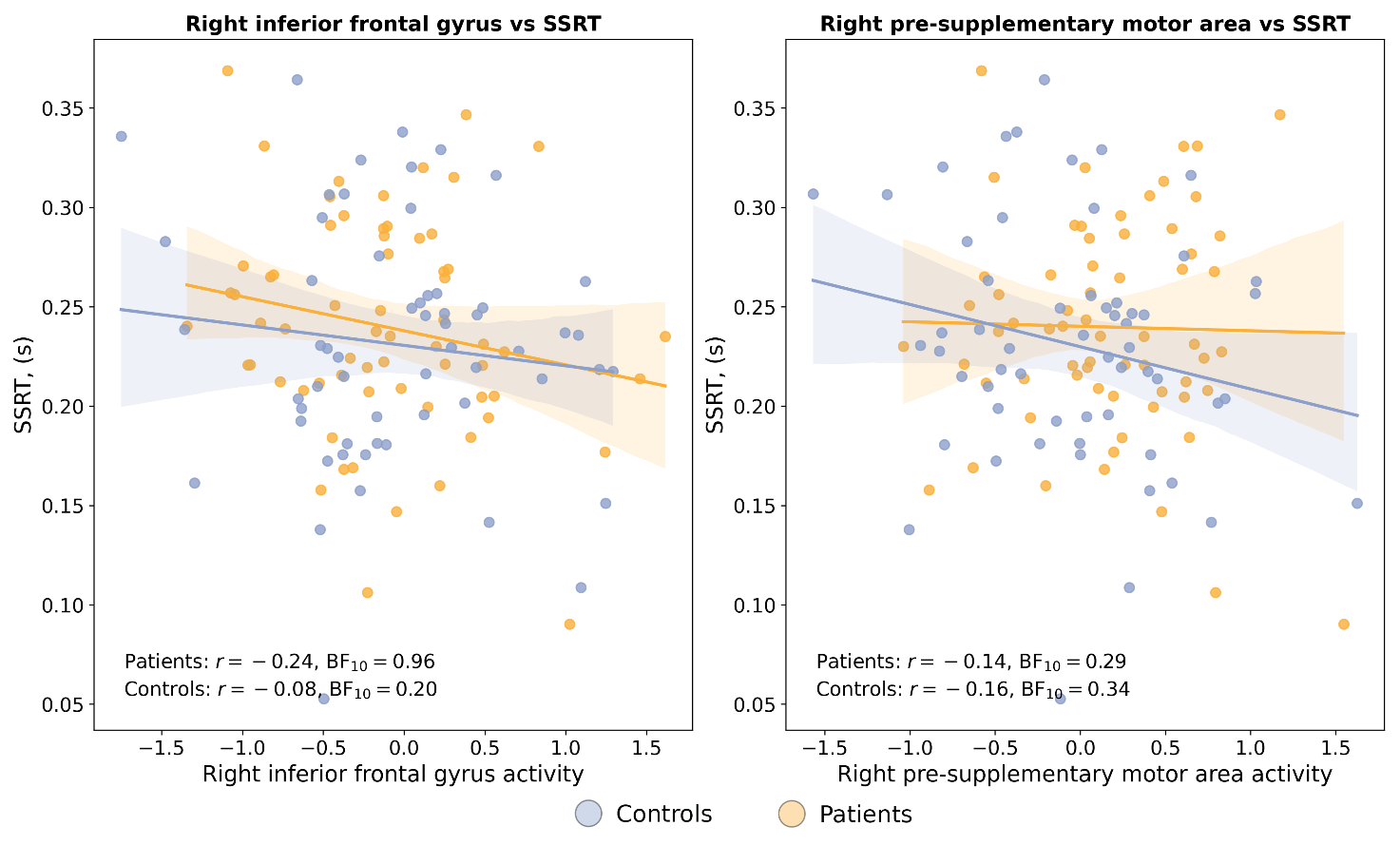

Figure S3: Association between stop-signal reaction time (SSRT) during successful inhibition within the right inferior frontal gyrus (left) and right pre-supplementary area (right) in patients (orange) and controls (blue).

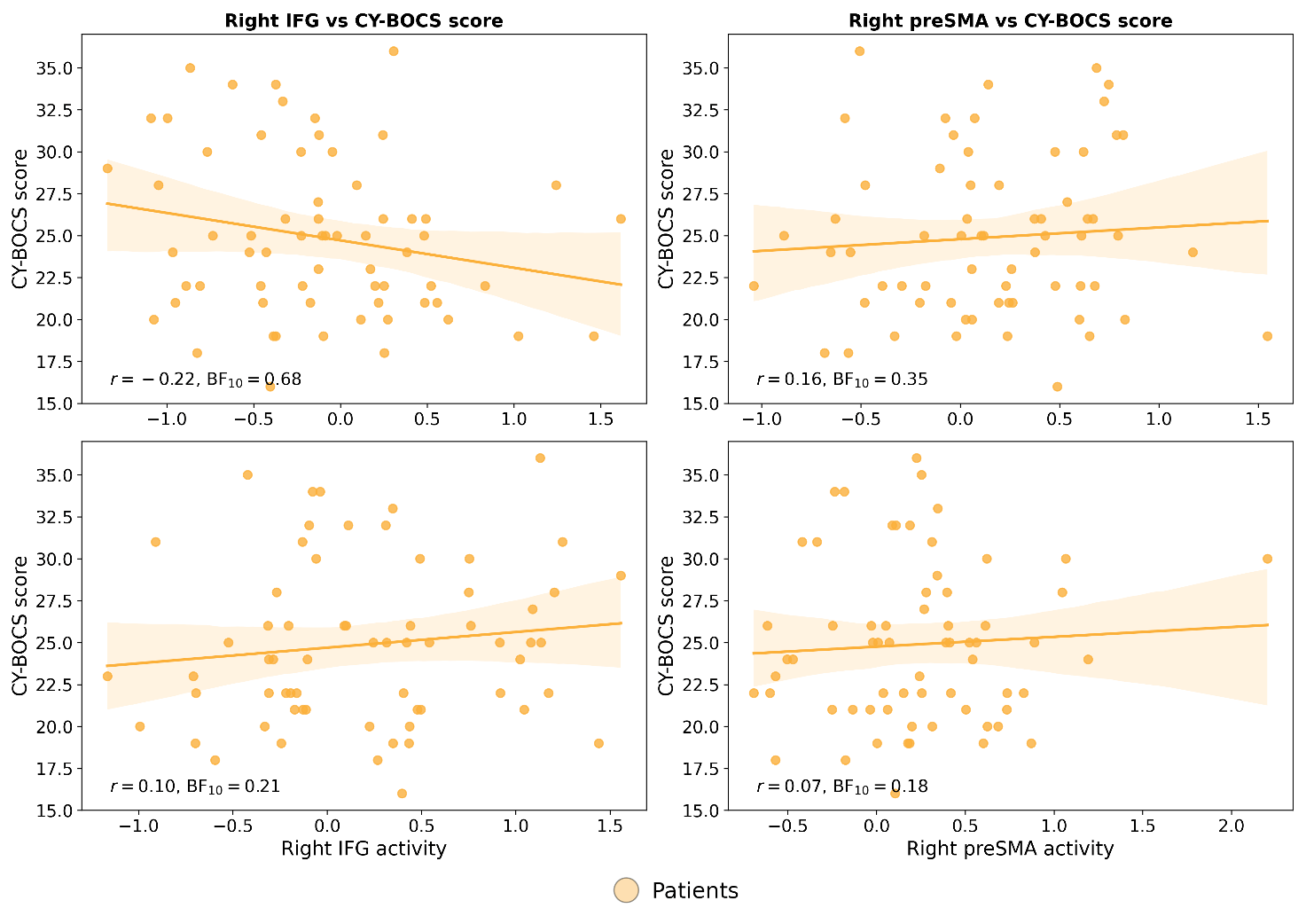

Figure S4: Association between CY-BOCS scores during successful inhibition (top) and failed inhibition (bottom) within the right inferior frontal gyrus (left) and right pre-supplementary area (right) in patients.
